# Secular Trends in Airway Management of Out-of-Hospital Cardiac Arrest in the National EMS Information System (NEMSIS) Dataset

**DOI:** 10.1101/2023.07.27.23293290

**Authors:** Christopher B. Gage, Jonathan R. Powell, Michelle Nassal, Henry Wang, Ashish R. Panchal

## Abstract

**Introduction:** Advanced airway management is essential in resuscitation from out-of-hospital cardiac arrest (OHCA). No longitudinal national studies have described longitudinal trends in airway device choice. We sought to evaluate secular trends of OHCA endotracheal intubation (ETI) and supraglottic airway (SGA) in the United States (US).

**Methods:** We evaluated ETI and SGA use for 2013-2022 in adult OHCA in the US using the National EMS Information System (NEMSIS) database. We identified OHCA events (CPR performed or defibrillation) and evaluated the proportions of ETI and SGA used during OHCA. We repeated the results stratified by urbanicity. We used descriptive statistics to describe incidence prevalence with nonparametric trend testing for proportional changes over time and a two-sample stochastic rank sum test for equivalence to evaluate airway use differences by urbanicity.

**Results:** During the study period, we observed 320,154,097 adult 9-1-1 events. Of 3,118,703 OHCA, there were 699,568 and 337,458 cases with reported ETI and SGA attempts. The dominant airway choice was ETI, though the trend of ETI choice decreased as SGA increased over time (p-trend <0.05). From 2013 to 2022, SGA use increased in urban settings while rural and suburban remained stable (urban 27% to 39%*; suburban 31% to 29%; rural 28% to 29%, respectively, *p<0.05).

**Conclusion:** Over ten years, rates of advanced airway use have increased, with ETI remaining the predominant airway for adults in OHCA. Interestingly, ETI choice decreased as SGA increased over the study period. SGA use distinctly differed in urban settings, increasing concerns for disparities in care provision among communities. With the increased use of SGA over time, further evaluation of patient outcomes is required in datasets with robust linkage to Utstein variables.

## Introduction

Approximately 350,000 people annually experience out-of-hospital cardiac arrest (OHCA), of which only 15% survive.^1^ Survival from OHCA depends on factors like bystander CPR, early defibrillation, and effective airway management. Prehospital airway management in OHCA care is essential, allowing for effective ventilation and optimizing resuscitation during cardiac arrest.^2^ ^3^ Prehospital clinicians perform airway management through differing combinations of non-invasive and invasive techniques, including bag-valve mask-only (BVM), supraglottic airway devices (SGA), and endotracheal intubation (ETI).

For decades ETI was the dominant advanced airway approach for OHCA. However, recent studies have promoted the utility of SGA in OHCA. Recent randomized control trial (RCT) data demonstrated that 72-hour survival, return of spontaneous circulation, hospital survival, and favorable neurological status at discharge are higher for SGA than ETI.^4^ SGA was more likely to have higher overall success rates, fewer first attempts, and fewer complications (e.g., unrecognized airway misplacement and inadequate ventilation) than ETI.^4^ Another RCT found that SGA was significantly more successful in achieving ventilation after up to 2 attempts, especially when SGA was placed before an ETI attempt.^5^ Even with the data described, the overall picture of airway management research in OHCA is limited.^6^

No national longitudinal study has evaluated whether the choice of advanced airway management between SGA and ETI in OHCA has changed. We sought to assess whether the proportions of these advanced airway devices have changed in OHCA patients over the last decade.

## Methods

### Study Design and Population

This retrospective study assessed airway device use in OHCA patients from the National Emergency Medical Services Information System (NEMSIS) dataset from 2013 to 2022. The American Institutes of Research Institutional Review Board approved the study.

The National Emergency Medical Services Information System (NEMSIS) is the national data standard for emergency medical services (EMS) in the United States. It was developed by the National Association of State EMS Directors and the National Highway Traffic Safety Administration in 1996. NEMSIS is designed to improve the quality and efficiency of EMS by providing a common language for exchanging data between EMS agencies and organizations and is used for various purposes, including quality improvement, research, and policy development.^7^ NEMSIS versions include Version 1 from 1996-2009, Version 2 from 2010-2016, and its current Version 3 starting in 2017.

### Selection of Subjects and Identification of Cases

We included NEMSIS 911 events for adults ≥18 years. Using similar methods developed by Chan et al. for the NEMSIS data set, we identified OHCA episodes using four constructs: 1) performance of CPR, 2) provision of CPR before or after EMS arrival, 3) chest compressions procedure, and 4) defibrillator or cardioversion procedure.^8^ Because NEMSIS cannot distinguish persons among the recorded events, we assumed each event represented a separate patient.

In 2017, NEMSIS implemented a transition from Version 2 to the Version 3 data standard. A salient update in Version 3 was the addition of the International Classification of Diseases (ICD) codes, ICD-9 and 10, now the coding standard for electronic healthcare records.^9^ Since this longitudinal analysis bridged the transition between data set versions, we used the NEMSIS crosswalk corresponding variable changes between the Versions 2 and 3 data sets.^10^ This crosswalk allows users to understand updates between variable identifiers from data dictionary 2.2.1 to 3.4.0, but unfortunately, it lists only data elements from Version 2 that remain in Version 3. Therefore, in addition to the crosswalk, we used Diggs 2014 and Wang 2011 for Version 2 (Appendix 1) and SNOMED CT Browser and Hanlin 2022 for Version 3 (Appendix 2) to categorize the different airway devices used in our OHCA population.^11–14^ The extracted variables for the different NEMSIS versions are noted in Appendix 1 (Version 2) and Appendix 2 (Version 3).

### Outcomes

The primary outcomes were the adult OHCA proportions receiving SGA or ETI. (Appendix 2) We did not include bag-value mask (BVM) ventilation because of inconsistent reporting in the NEMSIS data set. In addition, we did not include surgical airways because of their uncommon use in the OHCA.

### Measures

Demographic characteristics related to the analysis included age (year), gender (designated as male or female), and race (defined as White, Black or African American, Asian, American Indian or Alaska Native, Native Hawaiian or Other Pacific Island, or Other). Ethnicity was reported with race in Version 2 but separately in Version 3 (Hispanic/Latino and Not Hispanic/Latino). There were 246 ICD-10 codes for incident locations in NEMSIS (Appendix 3). These codes were categorized into Home/Residence, Healthcare Facility, Non-Healthcare Business, Street or Highway, and Other (e.g., sporting event, outdoors). Due to differences between versions, all variable codes are listed in their respective demographic tables.

The NEMSIS Project classified population setting (urbanicity) using the United States Department of Agriculture (USDA) and Office of Management and Budget (OMB) definitions: Urban (Urban Influence Codes 1, 2), counties with large (1+ million residents) or small (less than 1 million residents) metropolitan areas; Suburban (Urban Influence Codes 3 and 5), micropolitan (with an urban core of at least 10,000 residents) counties adjacent to a large or small metropolitan county; Rural (Urban Influence Codes 4, 6, 8, 9), non-urban core counties neighboring a large metropolitan area or a small metropolitan area (with or without a town); Wilderness (Urban Influence Codes 7, 10, 11, 12), non-core counties that are adjacent to micropolitan counties (with or without own town).^12, 15^ We used these classifications to evaluate airway proportions by urbanicity. Due to low population numbers, Wilderness populations were combined with Rural populations.^16^

### Statistical Analysis

We determined the annual incidence of each airway (ETI and SGA) and tested trends over time using a non-parametric test.^17^ We calculated the yearly advanced airway incidence rate by dividing the number of SGA by the total number of SGA plus ETI and the number of ETI divided by the total number of SGA plus ETI. We further stratified the analysis by urbanicity and used a two-sample stochastic rank sum test (TOST) for equivalence to assess for significant differences in SGA attempts among urbanicity between the years 2013 and 2022.^18^ Additionally, we evaluated the annual trend for the number of advanced airway devices per OHCA event. Descriptive statistics of population demographics were calculated. All analyses were conducted with Stata 17/Standard Edition.^19^

## Results

In the study period, there were 320,154,097 EMS activations reported in the NEMSIS data over the ten years, with 3,118,703 identified as adults suffering from OHCA (Figure 1). An average of 1.0% annually met the OHCA inclusion criteria in Version 2 and 0.92% in Version 3 (Table 1). The median age was similar over the study period, with males the most common gender (59.3%). There was a high amount of missingness for race (34.9%). OHCA events were more common in Urban (83.4%) and Home/Residential settings (65.2%) (Table 2).

**Figure 1:**
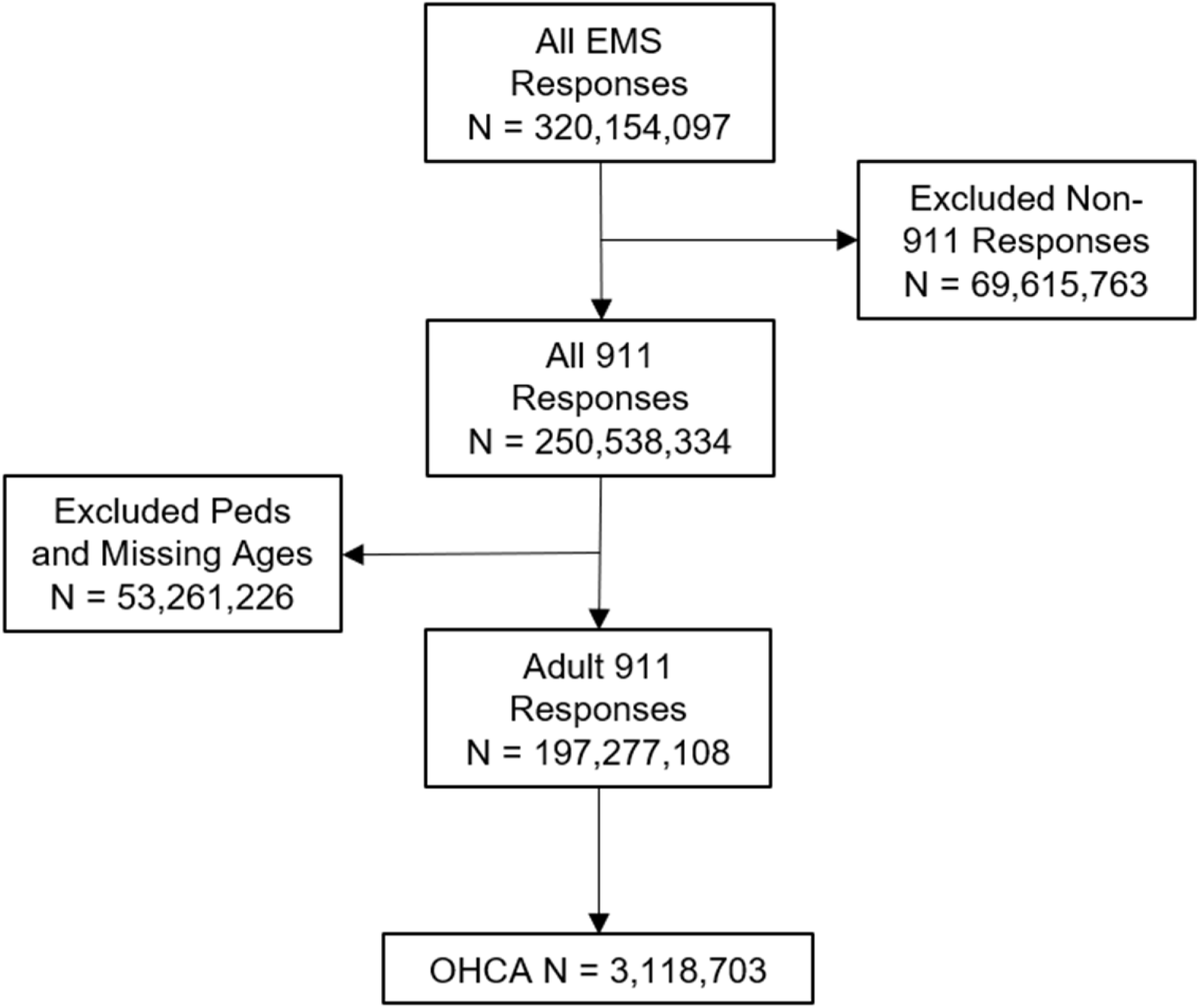
Study Design with Inclusion and Exclusion Criteria. Abbreviations: EMS – emergency medical services, OHCA – out-of-hospital cardiac arrest, Peds – pediatrics.

**Table 1:** Yearly proportions for inclusions, exclusions, and percentage for OHCA / All NEMSIS Calls. Abbreviations: CI – confidence interval, NEMSIS – national emergency medical services information system, OHCA – out-of-hospital cardiac arrest, Peds – pediatrics. Version 2: 2013-2016, Version 3: 2017-2022.

|  | 2013 | 2014 | 2015 | 2016 | 2017 | 2018 | 2019 | 2020 | 2021 | 2022 |
| --- | --- | --- | --- | --- | --- | --- | --- | --- | --- | --- |
| All NEMSIS Calls | 23,897,211 | 25,835,729 | 30,206,450 | 29,919,652 | 7,907,829 | 22,532,890 | 34,203,087 | 43,488,767 | 48,982,990 | 53,179,492 |
| Exclude Non-911 Events | 5,341,048 | 5,623,484 | 6,689,361 | 6,250,969 | 1,562,727 | 4,690,575 | 7,701,119 | 9,694,880 | 10,879,838 | 11,181,762 |
| 911 Responses | 18,556,163 | 20,212,245 | 23,517,089 | 23,668,683 | 6,345,102 | 17,842,315 | 26,501,968 | 33,793,887 | 38,103,152 | 41,997,730 |
| Exclude Peds and Missing | 3,814,650 | 4,096,026 | 4,805,988 | 4,988,810 | 1,272,537 | 3,995,932 | 5,762,803 | 7,182,452 | 8,191,589 | 9,150,439 |
| Adults and 911 Calls Only | 14,741,513 | 16,116,219 | 18,711,101 | 18,679,873 | 5,072,565 | 13,846,383 | 20,739,165 | 26,611,435 | 29,911,563 | 32,847,291 |
| Adults and 911 Calls Only (%) | 61.69% | 62.38% | 61.94% | 62.43% | 64.15% | 61.45% | 60.64% | 61.19% | 61.07% | 61.77% |
| OHCA Events | 267,999 | 258,961 | 287,712 | 328,745 | 68,745 | 197,933 | 287,028 | 437,284 | 481,142 | 503,154 |
| OHCA Events % (CI 95%) | 1.12 (1.12-1.13) | 1.00 (1.00-1.01) | 0.95 (0.95-0.96) | 1.10 (1.10-1.10) | 0.87 (0.87-0.88) | 0.88 (0.87-0.88) | 0.84 (0.84-0.84) | 1.01 (1.00-1.01) | 0.98 (0.98-0.99) | 0.95 (0.94-0.95) |

**Table 2:**
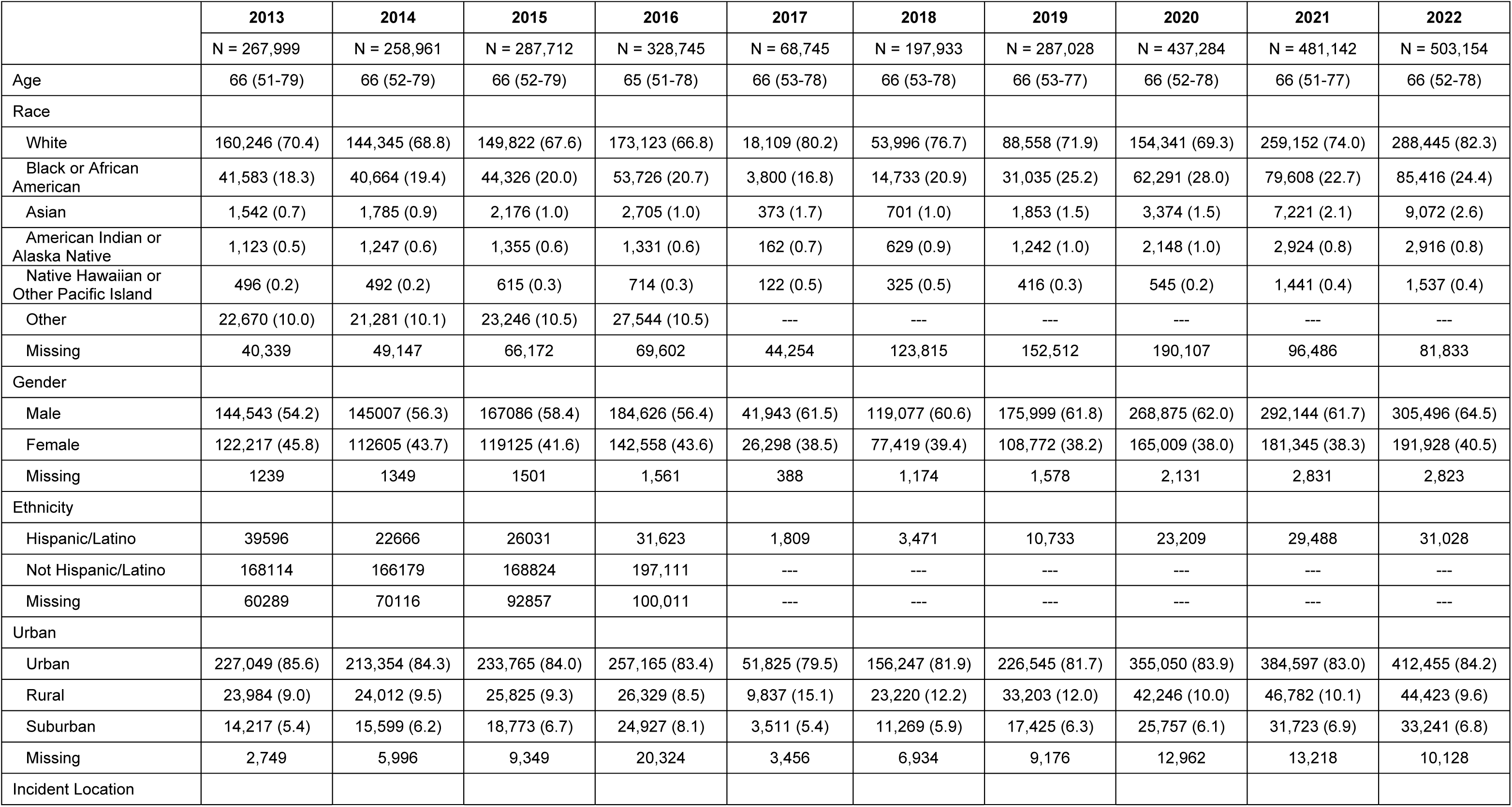
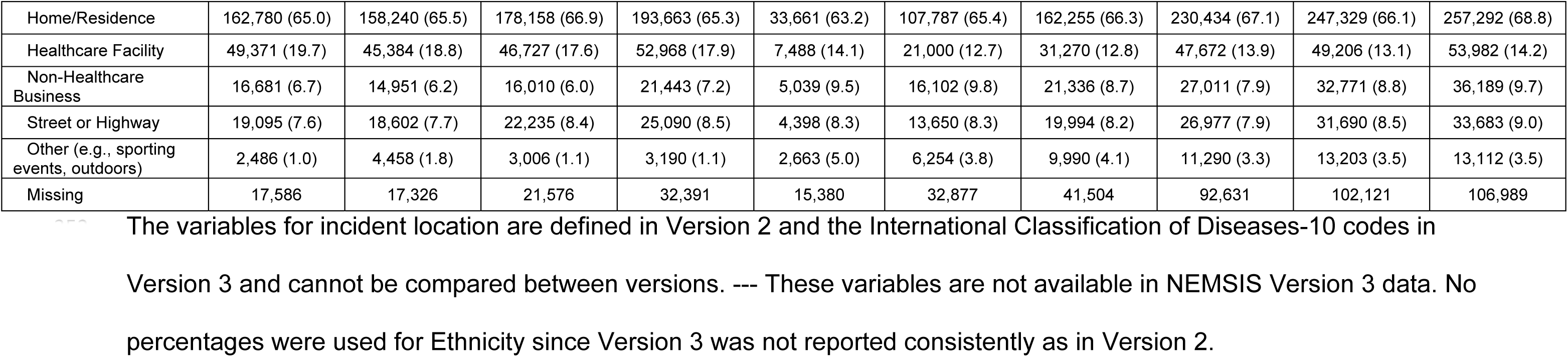
Demographics and characteristics (Frequency (%)) of out-of-hospital cardiac arrest patients and locations for National Emergency Medical Services Information System (NEMSIS) Version 2, 2013-2016, and Version 3, 2017-2022.

Over the last decade, the predominant airway approach used was ETI, and the percentage of advanced airways performed per OHCA events increased (Table 3). The proportional trend was significant (p-value <0.001) for SGA use increasing while the trend for ETI decreased (Figure 2). Specifically, ETI use appeared to decline over the last three years, with a subsequent increase in SGA use. When evaluated based on urbanicity (Figure 3), this trend was still true and appeared more focused in urban areas (e.g., decreased ETI use in urban areas with an associated increase in SGA).

**Figure 2:**
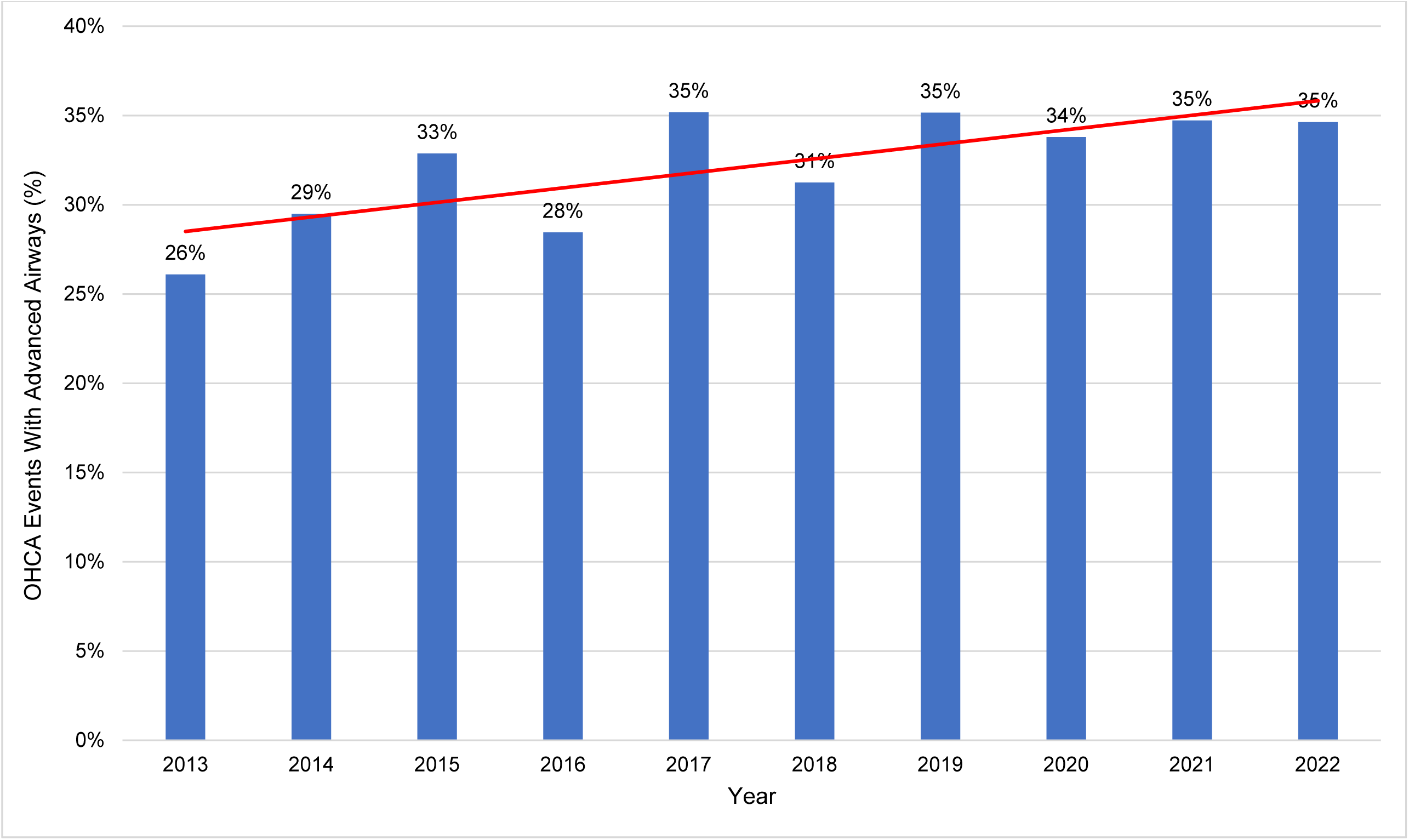
Advanced airway proportions per OHCA patient over ten years. Abbreviations: OHCA – out-of-hospital cardiac arrest.

**Figure 3:**
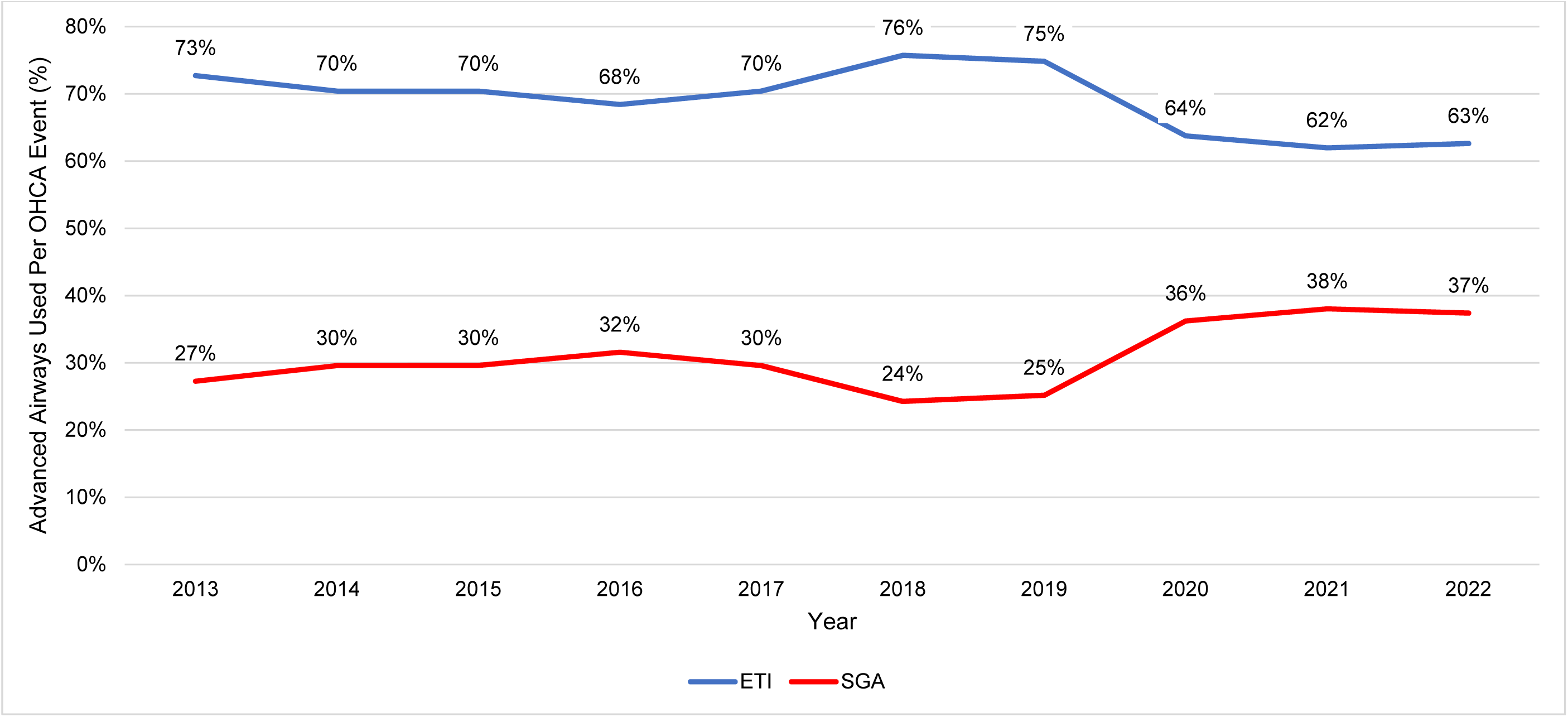
Percent of SGA and ETI use per total advanced airway events for OHCA over ten years. Abbreviations: ETI – endotracheal intubation, OHCA – out-of-hospital cardiac arrest, SGA – supraglottic airway.

**Figure 4:**
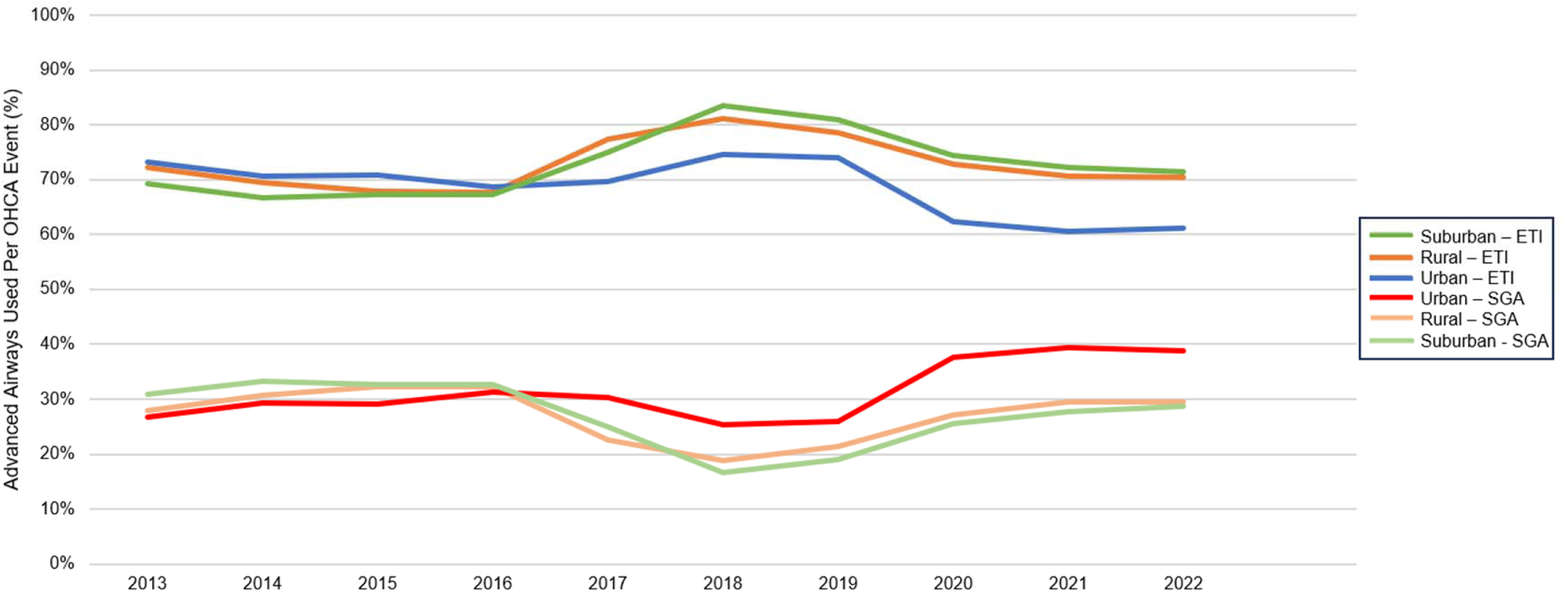
Overall SGA or ETI attempts for OHCA by urbanicity. Abbreviations: ETI – endotracheal intubation, OHCA – out-of-hospital cardiac arrest, SGA – supraglottic airway.

## Discussion

While advanced airway usage has increased in adult OHCA events, ETI usage has decreased and been replaced with SGA (Figure 2). This trend was more dramatic in urban areas compared to suburban or rural settings (Figure 3). Many factors can impact these choices including which level of EMS clinician arrives on the scene first, the local and state protocols, and patient position and location.^20^ Additionally, factors such as patient improvement, transportation time to the hospital, and the number of assisting responders, play a role in whether the initial device is maintained or if one switches to another.

Multiple RCTs have evaluated the impact of SGA first as a management strategy for OHCA, with one study demonstrating no difference in outcomes compared to ETI, while another demonstrated improved outcomes utilizing SGA.^4, 5^ Complications with ETI placement during OHCA often involve interruptions in chest compressions to allow for better vocal cord visualization that may lead to worsened return of spontaneous circulation (ROSC), and neurological outcomes.^21, 22^ Although there have been many advances around ETI, including gum elastic bougies and video laryngoscopy, concerns around reduced airway opportunities and lack of training may lead many EMS clinicians to choose the least invasive approach to airway management. Based on these and other evaluations, the 2020 American Heart Association (AHA) advanced cardiac life support (ACLS) guidelines recommend that ETI or SGA be used as primary advanced airway strategies for OHCA if ETI placement success rates are high.^23^ In situations where ETI placement success rates were low, the recommendation was to place SGA devices due to the higher first-pass success.

Our study demonstrates that over the last decade, SGA has been adopted by EMS clinicians as a valid advanced airway strategy. When SGA and ETI proportions are stratified by urbanicity (Figure 3), the degree of trend between ETI and SGA in urban settings differs in non-urban areas. The differences between urban and rural settings are noteworthy and could be multifactorial. First, urban and rural communities are different clinical practice settings. OHCA patients in non-urban regions tend to be in homes, have extended downtimes, and are unwitnessed.^24^ Rural areas tend to have more volunteer EMS clinicians, longer response and transport times, and sicker patients.^25^ Secondly, medical direction and knowledge dissemination practices may differ with urban areas having medical directors from larger, university-based medical centers that are more proactive in spreading current evidence-based knowledge.^25^ Differences in airway choice could be a factor leading to urban settings experiencing greater OHCA survival than those in non-urban settings, but a direct impact on patient outcomes is yet to be determined.^26^

Understanding the trends in airway management by urbanicity is a significant step in better understanding disparities in care throughout US communities. Linking airway device choice with Utstein variables would help determined the direct impact these decisions have on patient outcomes. Even without this type of robust evaluation, medical directors should consider enhancing dissemination and training strategies, especially in non-urban areas, to ensure high-level patient outcomes.

### Limitations

First, being a longitudinal study, one of the challenges encountered was the need to crosswalk the variables for OHCA and advanced airway devices from NEMSIS Version 2 (2013-2016) to Version 3 (2017-2022). Since Version 3 started incorporating the International Classification of Diseases (ICD) 10 codes and featured many new variables, a true crosswalk for these was unavailable. To combat this problem, we first leveraged the data dictionary, followed by searching for ICD-10 codes, and used previous evaluations to understand the differences in the variables (Appendices 1-3). Second, we limited our evaluation to nationally required variables due to data reporting and could not evaluate cardiac arrest Utstein variables and their association with airway device usage (e.g., shockable vs. non-shockable rhythm and witnessed). Third, race is difficult to quantify in NEMSIS due to a large amount of missing or misclassification. Additionally, ethnicity was combined into a unified race variable in version 3 making a true crosswalk difficult (Table 3). Finally, all data entry was performed by EMS clinicians with the possibility of reporting bias.

## Conclusion

Over ten years, rates of advanced airway use have increased, with ETI remaining the predominant airway for adults in OHCA. Interestingly, SGA use distinctly differed in urban settings, increasing concerns for disparities in care provision among communities. With the increased use of SGA over time, further evaluation of patient outcomes is required in datasets with robust linkage to Utstein variables.

## Conflict of interest statement

The authors declare no financial or other conflicts of interest.

### Abbreviations

CPR: cardiopulmonary resuscitation
BVM: bag valve mask
EMS: emergency medical services
ETI: endotracheal intubation
ICD: International Classification of Diseases
IRB: international research board
NEMSIS: National Emergency Medical Services Information System
OHCA: out of hospital cardiac arrest
ROSC: return of spontaneous circulation
SGA: supraglottic airway
SNOMED: systematized nomenclature of medicine

## Data Availability

All data in the manuscript is public access from the National EMS Information System (NEMSIS).

https://nemsis.org/

## Acknowledgments

We want to thank the NEMSIS Technical Assistance Center for their work in helping us access this data.

## Funding

This research received no specific grant from funding agencies in the public, commercial, or not-for-profit sectors.

## Notes

### Competing Interest Statement

The authors have declared no competing interest.

### Clinical Trial

N/A

### Funding Statement

No funding was used for this study.

### Author Declarations

The American Institutes of Research Institutional Review Board approved the study.

## References

1. Tsao CW, Aday AW, Almarzooq ZI, et al. Heart Disease and Stroke Statistics— 2022 Update: A Report From the American Heart Association. Circulation 2022;145(8). DOI: 10.1161/cir.0000000000001052.

2. Editorial Board. Circulation 2020;142(16_suppl_2):S336-S336. DOI: doi:10.1161/CIR.0000000000000929.

3. Kitamura T, Kiyohara K, Sakai T, et al. Epidemiology and outcome of adult out-of-hospital cardiac arrest of non-cardiac origin in Osaka: a population-based study. BMJ Open 2014;4(12):e006462. DOI: 10.1136/bmjopen-2014-006462.

4. Wang HE, Schmicker RH, Daya MR, et al. Effect of a Strategy of Initial Laryngeal Tube Insertion vs Endotracheal Intubation on 72-Hour Survival in Adults With Out-of-Hospital Cardiac Arrest: A Randomized Clinical Trial. JAMA 2018;320(8):769–778. DOI: 10.1001/jama.2018.7044.

5. Benger JR, Kirby K, Black S, et al. Effect of a Strategy of a Supraglottic Airway Device vs Tracheal Intubation During Out-of-Hospital Cardiac Arrest on Functional Outcome: The AIRWAYS-2 Randomized Clinical Trial. Jama 2018;320(8):779–791. (In eng). DOI: 10.1001/jama.2018.11597.

6. Agency for Healthcare Research and Quality (AHRQ). Guidelines and Measures Updates. September 2018 (https://www.ahrq.gov/gam/updates/index.html).

7. National EMS Information System (NEMSIS). What is NEMSIS. (https://nemsis.org/what-is-nemsis/).

8. Chan HK, Okubo M, Callaway CW, Mann NC, Wang HE. Characteristics of adult out-of-hospital cardiac arrest in the National Emergency Medical Services Information System. Journal of the American College of Emergency Physicians Open 2020;1(4):445–452. DOI: 10.1002/emp2.12106.

9. National Center for Health Statistics. International Classification of Diseases, (ICD-10-CM/PCS) Transition – Background. (https://www.cdc.gov/nchs/icd/icd10cm_pcs_background.htm#:~:text=CM%2FPCS%20Transition-,International%20Classification%20of%20Diseases%2C%20(ICD%2D10%2DCM%2F,Print).

10. National EMS Information System (NEMSIS). NEMSIS Version 2 to Version 3 Translation. (https://nemsis.org/media/nemsis_v3/release-3.4.0/Translation/v2v3Translation/v2v3Translation/NEMSIS_V2_V3_Translation_Data_Dictionary.pdf).

11. Diggs LA, Yusuf JE, De Leo G. An update on out-of-hospital airway management practices in the United States. Resuscitation 2014;85(7):885–92. (In eng). DOI: 10.1016/j.resuscitation.2014.02.032.

12. Wang HE, Mann NC, Mears G, Jacobson K, Yealy DM. Out-of-hospital airway management in the United States. Resuscitation 2011;82(4):378–85. (In eng). DOI: 10.1016/j.resuscitation.2010.12.014.

13. International SNOMED CT. SNOMED CT Browser. (https://browser.ihtsdotools.org/multi-extension-search.html).

14. Hanlin ER, Chan HK, Hansen M, et al. Epidemiology of out-of-hospital pediatric airway management in the 2019 national emergency medical services information system data set. Resuscitation 2022;173:124–133. (In eng). DOI: 10.1016/j.resuscitation.2022.01.008.

15. US Department of Agriculture. Urban Influence Codes. (https://www.ers.usda.gov/data-products/urban-influence-codes/).

16. Peters GA, Ordoobadi AJ, Panchal AR, Cash RE. Differences in Out-of-Hospital Cardiac Arrest Management and Outcomes across Urban, Suburban, and Rural Settings. Prehosp Emerg Care 2022:1–8. (In eng). DOI: 10.1080/10903127.2021.2018076.

17. Cuzick J. A Wilcoxon-type test for trend. Stat Med 1985;4(1):87–90. (In eng). DOI: 10.1002/sim.4780040112.

18. Lakens D. Equivalence Tests: A Practical Primer for t Tests, Correlations, and Meta-Analyses. Soc Psychol Personal Sci 2017;8(4):355–362. (In eng). DOI: 10.1177/1948550617697177.

19. Stata. Stata. (https://www.stata.com/).

20. National Association of State EMS Officials. National EMS Scope of Practice Model 2019 (Report No. DOT HS 812-666). National Highway Traffic Safety Administration. (https://www.ems.gov/pdf/National_EMS_Scope_of_Practice_Model_2019.pdf).

21. Grunau B, Kime N, Leroux B, et al. Association of Intra-arrest Transport vs Continued On-Scene Resuscitation With Survival to Hospital Discharge Among Patients With Out-of-Hospital Cardiac Arrest. JAMA 2020;324(11):1058–1067. DOI: 10.1001/jama.2020.14185.

22. Buick JE, Drennan IR, Scales DC, et al. Improving Temporal Trends in Survival and Neurological Outcomes After Out-of-Hospital Cardiac Arrest. Circ Cardiovasc Qual Outcomes 2018;11(1):e003561. (In eng). DOI: 10.1161/circoutcomes.117.003561.

23. Panchal AR, Bartos JA, Cabañas JG, et al. Part 3: Adult Basic and Advanced Life Support: 2020 American Heart Association Guidelines for Cardiopulmonary Resuscitation and Emergency Cardiovascular Care. Circulation 2020;142(16_suppl_2):S366-s468. (In eng). DOI: 10.1161/cir.0000000000000916.

24. Peters GA, Ordoobadi AJ, Panchal AR, Cash RE. Differences in Out-of-Hospital Cardiac Arrest Management and Outcomes across Urban, Suburban, and Rural Settings. Prehosp Emerg Care 2023;27(2):162–169. (In eng). DOI: 10.1080/10903127.2021.2018076.

25. King NP, Marcus; Huling, Sarah; Hanson, Brian EMS Services in Rural America: Challenges and Opportunities. National Rural Health Association: May 11, 2018 2018. (https://www.ruralhealth.us/NRHA/media/Emerge_NRHA/Advocacy/Policy%20documents/05-11-18-NRHA-Policy-EMS.pdf).

26. Kragholm K, Hansen CM, Dupre ME, et al. Care and outcomes of urban and non-urban out-of-hospital cardiac arrest patients during the HeartRescue Project in Washington state and North Carolina. Resuscitation 2020;152:5–15. (In eng). DOI: 10.1016/j.resuscitation.2020.04.030.

